# Social contact patterns among employees in 3 U.S. companies during early phases of the COVID-19 pandemic, April to June 2020

**DOI:** 10.1101/2020.10.14.20212423

**Authors:** Moses C. Kiti, Obianuju G. Aguolu, Carol Y. Liu, Ana R. Mesa, Rachel Regina, Kathryn Willebrand, Chandra Couzens, Tilman Bartelsmeyer, Kristin Nelson, Samuel Jenness, Steven Riley, Alessia Melegaro, Faruque Ahmed, Fauzia Malik, Ben Lopman, Saad B. Omer

## Abstract

2.

**Importance:** Devising control strategies against diseases such as COVID-19 require understanding of contextual social mixing and contact patterns. There has been no standardized multi-site social contact study conducted in workplace settings in the United States that can be used to broadly inform pandemic preparedness policy in these settings.

**Objective:** The study aimed to characterize the patterns of social contacts and mixing across workplace environments, including on-site or when teleworking.

**Design:** This was a cross-sectional non-probability survey that used standardized social contact diaries to collect data. Employees were requested to record their physical and non-physical contacts in a diary over two consecutive days, documented at the end of each day. Employees from each company were enrolled through email and electronic diaries sent as individual links. Data were collected from April to June 2020.

**Setting:** Two multinational consulting companies and one university administrative department, all located in Georgia, USA.

**Participants:** Employees opted into the study by accepting the invitation on a link sent via email.

**Main Outcome:** The outcome was median number of contacts per person per day. This was stratified by day of data collection, age, sex, race and ethnicity.

**Results:** Of 3,835 employees approached, 357 (9.3%) completed the first day of contact diary of which 304 completed both days of contact diary. There was a median of 2 contacts (IQR: 1-4, range: 0-21) per respondent on both day one and two. The majority (55%) of contacts involved conversation only, occurred at home (64%), and cumulatively lasted more than 4 hours (38%). Most contacts were repeated, and within same age groups, though participants aged 30-59 years reported substantial inter-generational mixing with children.

**Conclusion:** Participating employees in 3 surveyed workplaces reported few contacts, similar to studies from the UK and China when shelter-in-place orders were in effect during the pandemic. Many contacts were repeated which may limit the spread of infection. Future rounds are planned to assess changes in contact patterns when employees resume work in the office after the lockdown due to COVID-19 pandemic.

## 3. Introduction

The workplace is a key setting for social interactions and respiratory infection transmission, where 16% (range 9-33%) of influenza transmission is estimated to occur ^1^. Accordingly, the workplace is an important target to reduce severe acute respiratory syndrome coronavirus 2 (SARS-CoV-2) transmission through policies including remote work. In mid-April 2020, 62% of employed adults reported working remotely ^2^ in response to the COVID-19 pandemic. The potential impact of remote work on COVID-19 transmission can be quantified by assessing changes in social contact patterns ^3,4^. Mathematical models used to forecast epidemics and simulate the effects of interventions are highly sensitive to assumptions about who acquires infection from whom. These assumptions are based on data of social contact patterns, or the frequency and nature of contacts that individuals make on a daily basis.

There is a dearth of social contact data for the United States, especially in workplaces. One published telephone study from one US ^5^ state reported variation by age, day of the week and holiday period. Along with households, adults spend most of their time at work ^6^ where 20-25% of their contacts are with individuals from diverse ages and home locations ^1^. Our study aimed to characterize contact patterns relevant to respiratory infection spread during the COVID-19 pandemic among employees of two multinational consulting companies (N_1_=275, N_2_=3000) and one administrative department of a university (N_3_=560) in Atlanta, Georgia, from April to June 2020.

## 4. Methods

Company officials sent emails inviting all staff working in their US offices to participate. Participants opted into the study by consenting online. Each participant completed an enrolment survey which collected information on socio-demographics, the workplace, and household structure. Participants were expected to complete two consecutive days of online diaries through a personalized emailed survey link. Multiple daily contacts with the same person were recorded once, as well as the total time spent with that person. Contact with the same person on both days was counted as two contacts. A contact was defined as either non-physical (a two-way conversation with three or more words exchanged in the physical presence of another person) or physical (directly touching someone, either skin-to-skin or over the clothes).^7^ Remote work was defined as working from a location outside their designated site of work, including home or a public space. Each contact was characterized by age, sex, duration and location. For the second day of contact diary, we asked if each contact was repeated from the first day to assess contact persistence.

We summarized the median number of contacts per person per day stratified by age group, sex, race and household structure of the participant. We constructed age-specific contact matrices using four age groups for the participants (20-29, 30-39, 40-59, 60+ years) and six age groups for the contacts (0-9, 10-19, 20-29, 30-39, 40-59, 60+ years) and present age-specific average number of contacts per person per day. We assessed mixing within same age groups (assortative mixing) using the Q index (appendix 1) ^8^. All analysis was done in R v3.6.2. Ethical approval was given by Yale University (IRB# 2000026906).

## 5. Results

The overall response rate for the first day of the contact diary was 9.3% (357/3,835), of which 304 (85%) participants completed both days of contact diary. Because we found no substantial differences in characteristics between participants who completed one day only and both days, main results are summarized for the individuals who completed both.

The median age of the participants was 36 years (range 21-78) with around 50% aged 20-39 years and 10% older than 60 years (Table 1.) The majority self-identified as white (58%) and women (61%). Most participants reported at least a bachelor’s degree (94%), and all worked in office-based roles such as analysts, consultants, and managers. Household structures ranged from living with spouse only (32%), spouse and children only (25%), parent (8%), roommate or sibling (13%) or alone (15%); the most common state of residence was Georgia (49%). Almost all participants worked remotely on the day of diary completion (95%).

**Table 1.** Distribution of participant characteristics (n=304) and the median and interquartile range (IQR) of total contacts reported on the first diary day, April to June 2020.

|  | Total (%) | Median (IQR) Contacts -<br>First day | Daily Median (IQR)<br>Contacts – over two days |
| --- | --- | --- | --- |
| **Sex** |  |  |  |
| Women | 184 (60.5) | 2 (1-4) | 2 (1-4) |
| Men | 116 (38.2) | 2 (1-4) | 2 (1-4) |
| Prefer not to answer | 4 (1.3) | 0.5 (0-1.5) | 1 (0-1.5) |
| **Age Group (years)** |  |  |  |
| 20-29 | 90 (29.6) | 2 (1-3) | 2 (1-3) |
| 30-39 | 76 (25.0) | 2 (1-3) | 2 (1-3) |
| 40-49 | 60 (19.7) | 3 (2-5) | 3 (1-5) |
| 50-59 | 49 (16.1) | 2 (1-3) | 2 (1-3) |
| 60+ | 29 (9.5) | 2 (1-4) | 2 (1-4) |
| **Race** |  |  |  |
| Black | 26 (8.6) | 2.5 (1-4) | 2 (1-4) |
| White | 174 (57.2) | 2 (1-4) | 2 (1-4) |
| Asian | 48 (15.8) | 2 (0.8-3) | 1 (1-3) |
| Mixed | 52 (17.1) | 2 (1-4) | 2 (1-4) |
| Other | 4 (1.3) | 3 (1.8-4.3) | 3 (1-4) |
| **Hispanic or Latino** |  |  |  |
| Yes | 14 (4.6) | 3 (1.3-5) | 3 (1-4) |
| No | 290 (95.4) | 2 (1-4) | 2 (1-4) |
| **Household structure <sup>a</sup>** |  |  |  |
| Live alone | 44 (14.5) | 0.5 (0-2) | 0 (0-2) |
| Live with parent | 26 (8.6) | 3 (2-5.8) | 3 (2-5) |
| Roommate or sibling | 39 (12.8) | 3 (2-5) | 3 (1-4) |
| Spouse and children only | 76 (25.0) | 3 (2-5) | 3 (2-4) |
| Spouse only | 97 (31.9) | 1 (1-2) | 1 (1-2) |
| Other <sup>b</sup> | 22 (7.2) | 3 (2-4) | 2 (1-4) |

<sup>a</sup> Household structure was considered as living with parent if the participant lived with parent regardless of who else was in the house. This category includes households with parents, participant, spouse; parents, participant, children etc.
<sup>b</sup> Others are living with children only, grandchildren, grandparents, friends, niece, nephew, tenant and au pair.

A median of 2 contacts (IQR: 1-4, range: 0-21) was reported per respondent on both days. Over both days, a median of 2 contacts (IQR: 1-4) was reported. The number of contacts differed by household structure but not by sex, age group, race or ethnicity. Individuals living alone or living with their spouse only reported a median of 0 contacts (IQR: 0-2) which was lower than individuals living with spouse and children, sibling or roommate or with at least a parent (median 3; Table 1).

Of the 608 diary-days, 202 (33%) were collected in April and 406 (67%) were collected in May and June. Among the 1,548 social contacts reported across two days of data collection, 55% were conversation only, 12% were physical contact only, and 34% were conversation and physical contact (Figure 1). Among these, 64% occurred at home, 14% on the streets or at stores, 6% at work, and 5% at another person’s home (Panel A). We report both long lasting (38% of contacts lasting >4 hours) and short (32% lasting <15 minutes) contacts. 97% of long-lasting contacts occurred at the participant’s home while 78% of short contacts occurred outside. Sixty two percent of contacts were repeated over both days of data collection. 87% of repeat contacts occurred in the participant’s home.

**Figure 1.**
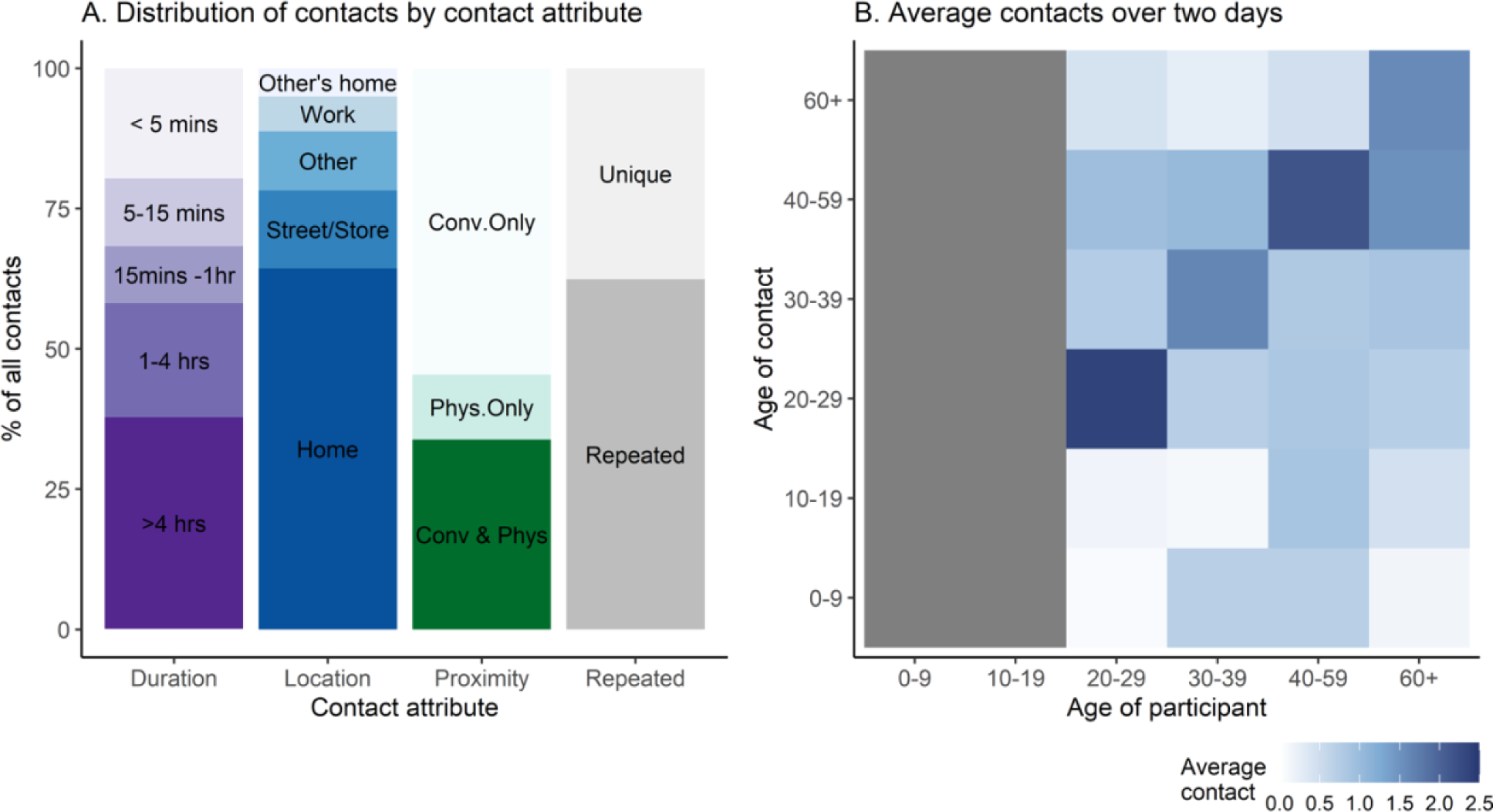
Panel (A) shows the distribution of contacts by attributes: duration (in minutes (mins) or hours (hr)). Types of contact were conversation with physical touch (Conv & Phys), physical only (Phys), or non-physical/conversation only (Conv only). A contact was repeated if observed on both days or unique if observed on only one day. Panel (B) shows the age-stratified average number of contacts over two study days. The gray area on the x-axis indicates that all respondents were over the age of 19, however they were able to report contacts under the age of 19 years. Data shown in the graphs are for 1,548 contacts recorded by 304 participants over 608 diary-days

The age-stratified contact matrix (Panel B) details average contacts made by participants and contacts within and across age groups. The peak mean numbers of contacts were on the main diagonal, indicating preferential mixing among individuals of the same age group. However, there was little evidence of assortative mixing by age for contacts above 20 years (Q index=0.27). The mixing matrix also shows inter-generational mixing involving 30-59-year-old participants and children.

## 6. Discussion

Participating study employees reported a median of 2 contacts from April to June 2020, when shelter-in-place orders were in effect in Georgia and most other states ^7^. Of the contacts that were reported, the majority were at home and were repeated rather than new. As such, employees who responded to the survey were not making many new contacts during this phase of the pandemic, which could limit transmission risk.

Recruitment in companies was online and elicited few responses despite regular reminders and monetary incentives. Employees interested in public health or who adhere to social distancing policies may be more likely to respond. Public health messaging calling for social distancing may have created stigma around reporting high contact rates resulting in the low contact rates reported in this study. These selection and information biases may have led to underreporting of contact rates. We lack pre-pandemic data from this specific population for direct comparison, but other studies in the UK ^10^ and China ^4^ report reduced contact rates by office workers during the pandemic. During the COVID-19 pandemic, many U.S. companies instituted work-from-home policies ^11^, likely drastically reducing total number of contacts in workplaces and the general population. In subsequent rounds of the study, we will assess changes in contact patterns as employees return to work in office settings. In future, we aim to use the contact patterns to parameterize mathematical models describing risk of disease transmission and investigate prevention and control strategies to prevent transmission in the workplace.

## Data Availability

All de-identified data and code used for analysis are available in Github through the link provided.

https://github.com/lopmanlab/corpmix

## 8. Supplementary Information

## Appendix 1. Calculation of Q-index for assortativeness

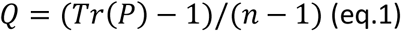

Where Tr(P) denotes the Trace of the matrix and P [*p*_*ij*_] is the matrix whose elements *p*_*ij*_ represent the proportion of total contacts of age group i with age group j. The elements are computed by the equation:

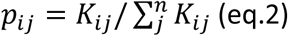

Where *K*_*ij*_ is the total contacts of age group i with age group j, 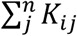 is the total contacts made by age group i and n is the total number of participant age groups.

The Q index ranges between negative one, corresponding to fully disassortative mixing, and one, under full diagonal dominance, i.e. fully assortative mixing. A Q index of zero corresponds to proportionate mixing. Therefore, Q represents a measure of departure from proportionate mixing for groups defined on a qualitative scale^8^.

## Appendix 2.

Distributions of percentage of contacts by location (A), duration (B), type (C) and persistence (D) based on participants’ age group. In panel C, contacts are classified as conversation only (Conv only), conversation with physical touch (Conv & Phys) and physical touch only (Phys only).

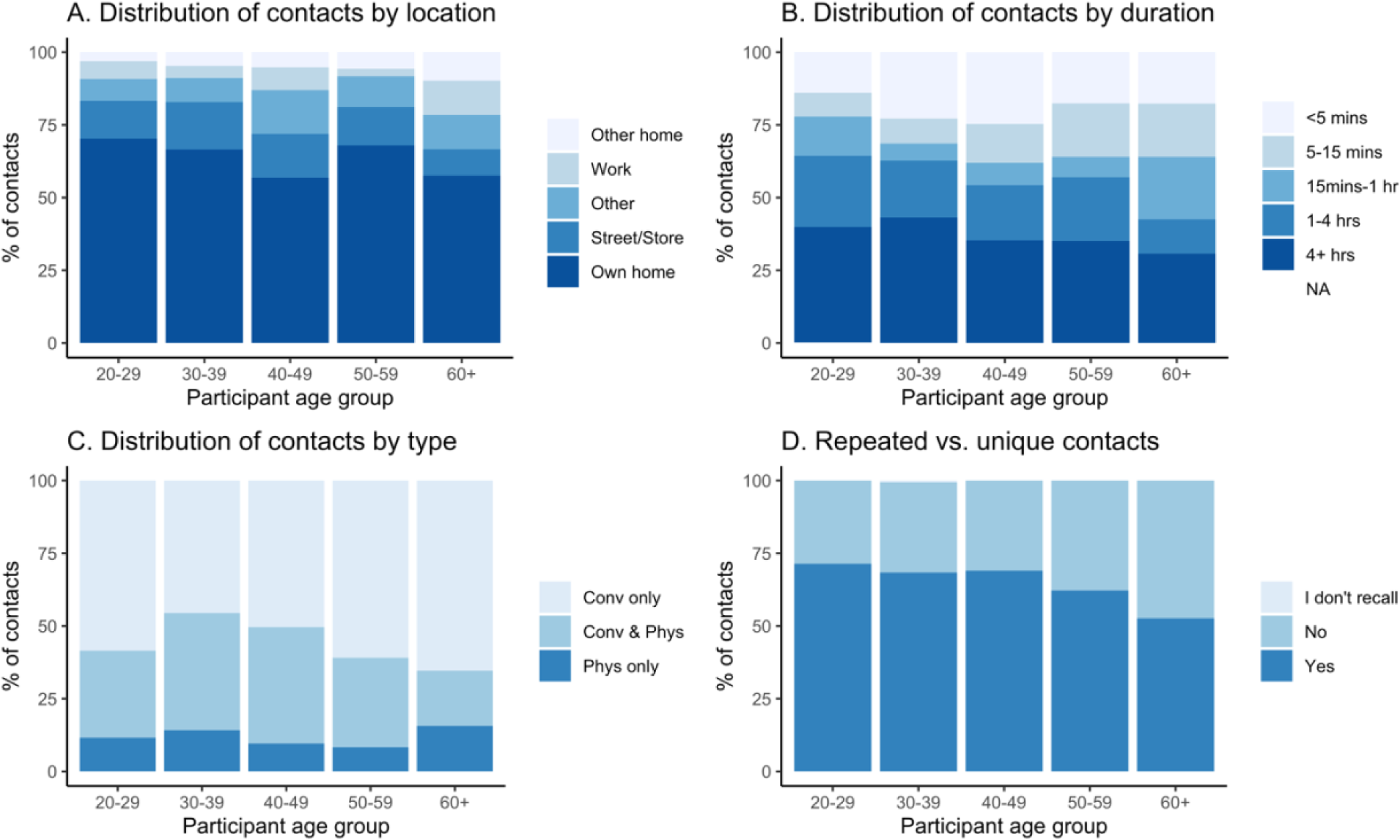

## Appendix 3.

Additional age-stratified mixing matrices by various contact attributes. Contact matrices at home, street or store, work, repeated over two days and unique over two days represent both conversation and physical contacts. In all panels, the grey area indicates that no data was collected from individuals aged 0-19 years.

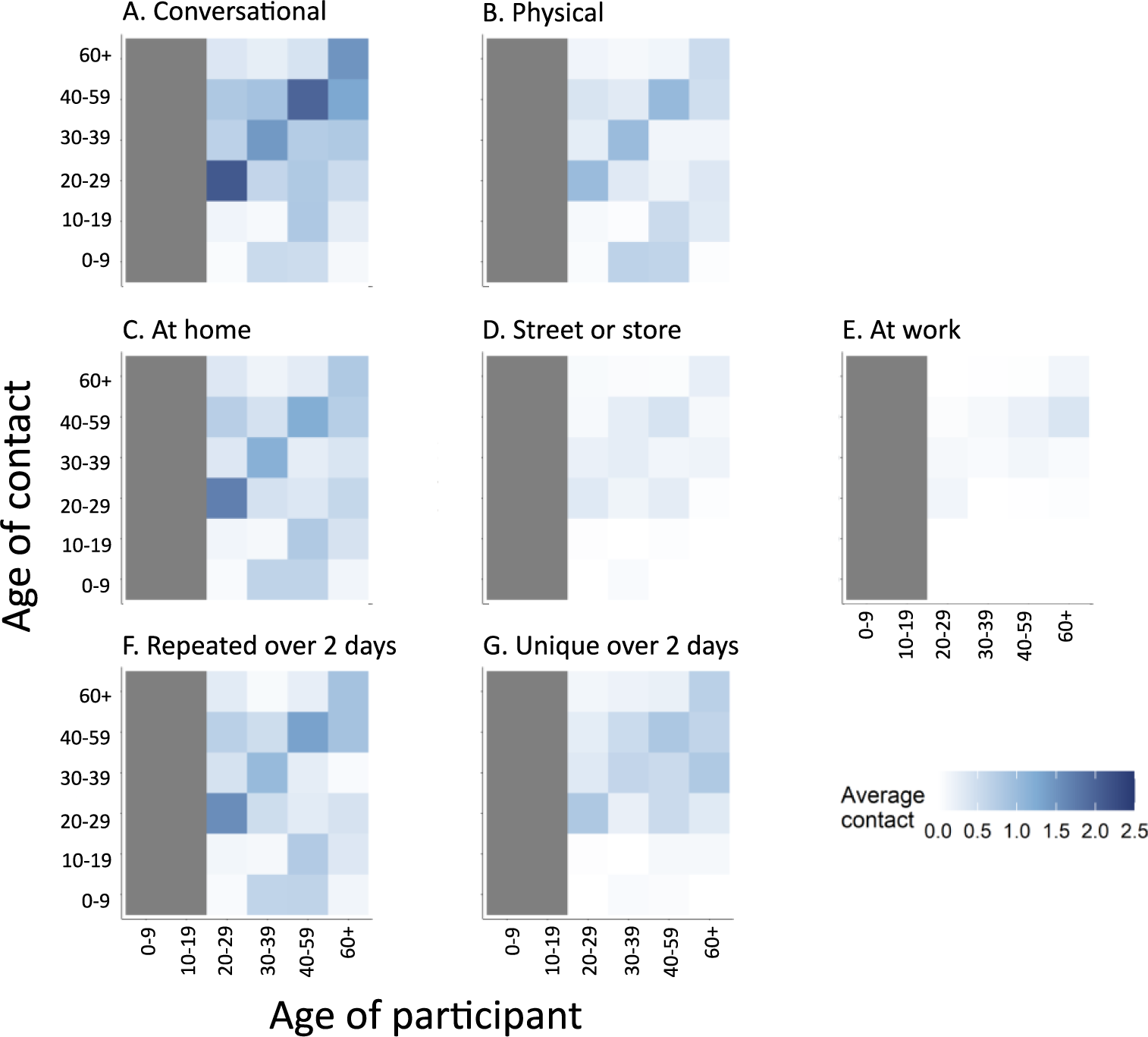

## Author Contributions

Dr. Saad and Dr. Lopman had full access to all of the data in the study and take responsibility for the integrity of the data and the accuracy of the data analysis.

*Concept and design:* Saad Omer, Ben Lopman, Obianuju Aguolu, Faruque Ahmed

*Acquisition, analysis, or interpretation of data:* Obianuju Aguolu, Ana Mesa, Katherine Willebrand, *Chandra Couzens*, Tilman Bartelsmeyer, Carol Liu, Moses Kiti

*Drafting of the manuscript:* Moses Kiti, Carol Liu, Kristin Nelson

*Critical revision of the manuscript for important intellectual content:* Saad Omer, Ben Lopman, Obianuju Aguolu, Kristin Nelson, Faruque Ahmed, Steven Riley, Alessia Melegaro

*Statistical analysis:* Carol Liu, Moses Kiti

*Administrative, technical, or material support:* Faruque Ahmed

*Supervision:* Saad Omer, Ben Lopman

## Additional Contributions

None

